# Determinants of COVID-19 Vaccine Acceptance in the U.S.

**DOI:** 10.1101/2020.05.22.20110700

**Authors:** Amyn A. Malik, SarahAnn M. McFadden, Jad Elharake, Saad B. Omer

**Affiliations:** Yale Institute for Global Health, New Haven, Connecticut, United States of America; Department of Internal Medicine, Infectious Disease, Yale School of Medicine, New Haven, Connecticut, United States of America; Yale School of Public Health, New Haven, Connecticut, United States of America; Yale School of Nursing, Orange, Connecticut, United States of America

**Keywords:** Covid-19, vaccine acceptance, evidence-based messaging, health disparitie

## Abstract

The COVID-19 pandemic continues to adversely affect the U.S., which leads globally in total cases and deaths. As COVID-19 vaccines are under development, public health officials and policymakers need to create strategic vaccine-acceptance messaging to effectively control the pandemic and prevent thousands of additional deaths.

**Methods:** Using an online platform, we surveyed the U.S. adult population in May 2020 to understand risk perceptions about the COVID-19 pandemic, acceptance of a COVID-19 vaccine, and trust in sources of information. These factors were compared across basic demographics.

**Findings:** Of the 672 participants surveyed, 450 (67%) said they would accept a COVID-19 vaccine if it is recommended for them. Males (72%), older adults (≥55 years; 78%), Asians (81%), and college and/or graduate degree holders (75%) were more likely to accept the vaccine. When comparing reported influenza vaccine uptake to reported acceptance of the COVID-19 vaccine: 1) participants who did not complete high school had a very low influenza vaccine uptake (10%), while 60% of the same group said they would accept the COVID-19 vaccine; 2) unemployed participants reported lower influenza uptake and lower COVID-19 vaccine acceptance when compared to those employed or retired; and, 3) black Americans reported lower influenza vaccine uptake and lower COVID-19 vaccine acceptance than nearly all other racial groups. Lastly, we identified geographic differences with Department of Health and Human Services regions 2 (New York) and 5 (Chicago) reporting less than 50 percent COVID-19 vaccine acceptance.

**Interpretation:** Although our study found a 67% acceptance of a COVID-19 vaccine, there were noticeable demographic and geographical disparities in vaccine acceptance. Before a COVID-19 vaccine is introduced to the U.S., public health officials and policymakers must prioritize effective COVID-19 vaccine-acceptance messaging for all Americans, especially those who are most vulnerable.

**Funding:** The study was funded by the Yale Institute for Global Health.

## Introduction

The coronavirus disease 2019 (COVID-19) pandemic has spread across the world with millions infected and hundreds of thousands dead.^1,2^ While most countries impacted have developed successful response strategies and observed significant improvements, the U.S. (as of May 21st, 2020) leads globally with 1·55 million cases and over 93,000 deaths.^3^ Additionally, according to the Centers for Disease Control and Prevention (CDC), current data show a disproportionate burden of COVID-19 infections and deaths among racial and ethnic minority communities.^4^ With the U.S. facing an economic disruption and the future remaining unknown, a vaccine to prevent COVID-19 infection is perhaps the best hope for ending the pandemic.

As misinformation about COVID-19 has spread across media outlets, it is important for U.S. public health officials and politicians to begin planning for effective messaging and policies before a vaccine is introduced. The U.S. already struggles with reaching high rates of influenza vaccine coverage—with less than half of the adult population vaccinated in 2019, resulting in over 30,000 vaccine-preventable deaths—therefore, COVID-19 presents an imminent danger that requires immediate action.^5^ Health communication must reach all communities, especially the most vulnerable, to educate Americans about the safety of vaccines and prevent future infections and deaths.

Immunization programs are only successful when there are high rates of acceptance and coverage. To accomplish this, it is critical to understand Americans’ risk perceptions about COVID-19, acceptance of a COVID-19 vaccine, and confidence in media sources, specifically those used to obtain information about the COVID-19 pandemic. The purpose of our study is to describe the current vaccine acceptance landscape with aims to 1) predict COVID-19 vaccine acceptance using regularly available demographic information, 2) identify the most vulnerable populations, and 3) provide information for public health officials and politicians to develop messaging for all Americans, while targeting communities most in need.

## Methods

Data were collected using an electronic questionnaire via Qualtrics® (Qualtrics, Provo, UT). In early May 2020, participants completed a questionnaire on CloudResearch.^6^ CloudResearch is an online survey platform that allows for representative surveying. The goal of our sampling was to be representative of the U.S. general population based on age, gender, education, race and ethnicity. Participants were eligible if they were 18 years of age or older, could read English, and had a CloudResearch account with access to the internet via computer or smart phone. Participants received compensation in the amount they agreed to with the platform through which they entered this survey; these rewards could include gift cards or donations to a participant selected charity.

Basic demographic information was collected as well as zip code, state of residence, and employment status. Additionally, we asked participants if a COVID-19 vaccine were available and recommended for them, would they would accept it (5-point Likert Scale: 1 = strongly disagree to 5 = strongly agree); this variable was dichotomized to COVID-19 vaccine acceptance (0 = strongly disagree/disagree/neutral; 1 = agree/strongly agree). Participants also completed the perceived risk scale (Cronbach’s *α* =0·70) which had 10 survey-items (5-point Likert Scale: 0 = strongly disagree/disagree/neutral; 1 = agree/strongly agree). Of the 672 participants, participants were removed from score calculation if they did not respond to one or more the of the items on the risk perception scale or selected “don’t know,” so this score was calculated for 537 participants. We also asked participants if they had received the influenza vaccine in the previous 8 months. Finally, participants were asked about their confidence in media sources and the reliability of these sources regarding the COVID-19 pandemic (5-point Likert Scale: 1 = strongly disagree to strongly agree). Yale University Institutional Review Board approved this study (IRB protocol number: 2000027891). Participants provided informed consent prior to data collection.

### Statistical Analysis

Descriptive statistics (frequencies, percentage) were calculated for the sample demographic characteristics. Additionally, the frequency and percentage of COVID-19 vaccine acceptance and reported influenza vaccination status were calculated. A chi-square analysis was completed to compare the reported influenza uptake to the reported COVID-19 vaccine acceptance.

We collapsed COVID-19 vaccine acceptance and influenza vaccine uptake into the 10 regions developed by the Department of Health and Human Services (DHHS). We then mapped these percentages onto a U.S. map. The 10 HHS regions are Region 1-Boston (Connecticut, Maine, Massachusetts, New Hampshire, Rhode Island, and Vermont), Region 2-New York (New Jersey, New York, Puerto Rico, and the Virgin Islands), Region 3-Philadelphia (Delaware, District of Columbia, Maryland, Pennsylvania, Virginia, and West Virginia), Region 4-Atlanta (Alabama, Florida, Georgia, Kentucky, Mississippi, North Carolina, South Carolina, and Tennessee), Region 5-Chicago (Illinois, Indiana, Minnesota, Ohio, and Wisconsin), Region 6-Dallas (Arkansas, Louisiana, New Mexico, Oklahoma, and Texas), Region 7-Kansas City (Iowa, Kansas, Missouri, and Nebraska), Region 8-Denver (Colorado, Montana, North Dakota, South Dakota, Utah, and Wyoming), Region 9-San Francisco (Arizona, California, Hawaii, Nevada, American Samoa, Commonwealth of the Northern Mariana Islands, Federated States of Micronesia, Guam, Marshall Islands, and Republic of Palau), and Region 10-Seattle (Alaska, Idaho, Oregon, and Washington).^7^

To assess the associations (odds ratios) of demographic factors with COVID-19 vaccine acceptance, a logistic regression analysis was used. Model selection using stepwise backward selection with a p-value of 0·2 was used to select the final, parsimonious model where age, gender, race, education, ethnicity, and employment status were included as explanatory variables. We computed area under the receiver operating curve (AUC) for our final model to evaluate model performance. We used bootstrap resampling (1000 samples) for internal validation and to obtain an area under the curve value accounting for model optimism.^8,9^ Data were analyzed using Stata version 16 (StataCorp, College Station, Texas).

## Results

We invited 2,010 participants, of which 672 (33%) completed the survey with 386 (57%) females and 256 (38%) 55 years old or over. The majority of the participants were non-Hispanic white (n = 436; 65%) and had a college or graduate degree (n = 351; 52%). The median risk perception score for COVID-19 was 6 (IQR: 5 – 7; mean: 5.9; SD: 2·0). Table 1 shows the demographics characteristics of the survey participants and the general U.S. population.

Of the 672 participants surveyed, 450 (67%) said they would accept a COVID-19 vaccine if it is recommended for them. The vaccine acceptance differed by demographic characteristics with males (72%), older people (55 years and above; 78%), Asian (81%), and college or graduate degree holders (75%) more likely to accept the vaccine if it would be recommended for them (table 1; figure 1). The median risk perception score amongst those who would accept the vaccine was 6 (IQR: 6 – 8) compared to a median of 5 (IQR: 2 – 6) amongst those who would not accept the vaccine. This difference in risk perception score was statistically significant (p < 0.01). As sensitivity analysis, we carried out a weighted analysis for COVID-19 vaccine acceptance using U.S. demographics.^10^ Weighing by age and sex decreased the percent acceptance to 62% while weighing by age, sex, and race decreased the percent acceptance to 57%.

Vaccine acceptance for COVID-19 also differed compared to the influenza vaccine with 348 (52%) of the participants having received the influenza vaccine in the last eight months (chi sqr p-value < 0·01; figure 1). Notable demographic differences exist when comparing reported influenza vaccine uptake to reported acceptance of the COVID-19 vaccine. For example, participants who did not complete high school, had a very low influenza vaccine uptake (n = 1; 10%), but of that same group, 60% (n = 6) said they would accept the COVID-19 vaccine if it were available and recommended for them. Another interesting demographic difference is that the part of the sample that reported being unemployed reported lower influenza uptake and lower COVID-19 vaccine acceptance when compared to those who reported being employed or retired. Additionally, black Americans reported lower influenza vaccine uptake (n = 28; 42%) and lower COVID-19 vaccine acceptance (n = 27; 40%) than nearly all other racial groups. A final demographic difference is that older adults reported higher influenza vaccine uptake (n = 177; 69%) and higher COVID-19 vaccine acceptance (n = 200; 78%) than younger participants.

**Figure 1.**
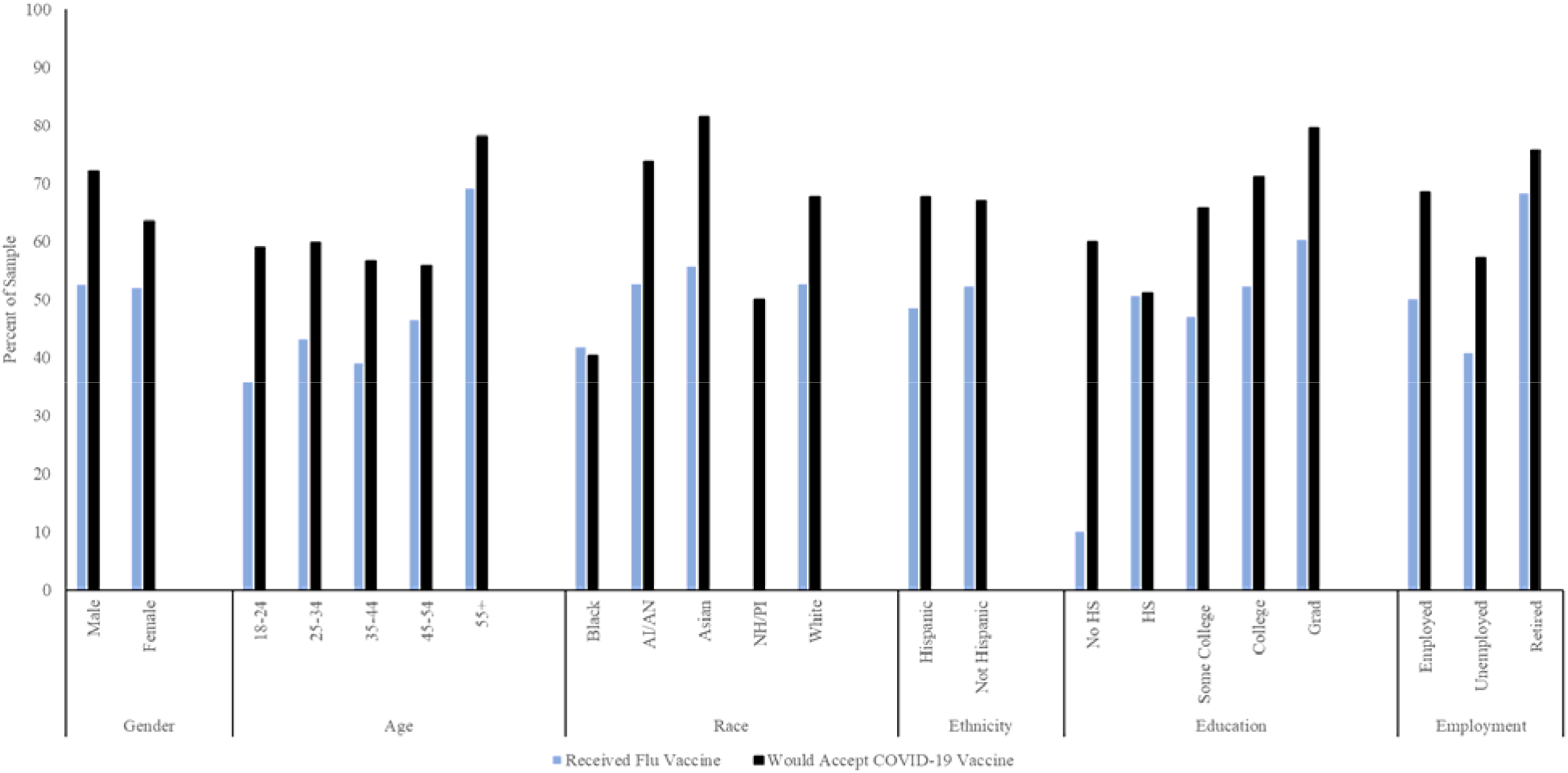
Comparison by demographic categories of the percent of the sample who reported receiving the influenza vaccine to those would reported they would accept the COVID-19 vaccine.

**Table 1.** Risk Perception and COVID-19 Vaccine Acceptance across Demographic Characteristics.

|  | **Total**<br>**(N = 672)**<br>**n (%)** | **Risk Perception Score**<br>**(out of 10; M +/- SD)** | **COVID-19**<br>**Vaccine**<br>**Acceptance**<br>**n (%)** | **U.S.**<br>**Population<sup>10</sup>**<br>**(%)** |
| --- | --- | --- | --- | --- |
| Gender |  |  |  |  |
| Male | 280 (42) | 5.9 +/- 2.0 | 202 (72) | 49 |
| Female | 386 (57) | 5.9 +/- 2.1 | 245 (63) | 51 |
| Other | 6 (1) | 3.8 +/- 3.0 | 3 (50) | 0.0 |
| Age (years) |  |  |  |  |
| 18-24 | 73 (11) | 5.4 +/- 2.2 | 43 (59) | 12 |
| 25-34 | 107 (16) | 5.5 +/- 2.0 | 64 (60) | 18 |
| 35-44 | 141 (21) | 5.9 +/- 2.0 | 90 (64) | 16 |
| 45-54 | 95 (14) | 5.4 +/- 2.0 | 53 (56) | 17 |
| 55+ | 256 (38) | 6.3 +/- 2.0 | 200 (78) | 36 |
| Race |  |  |  |  |
| Black/African American | 67 (10) | 5.4 +/- 2.1 | 27 (40) | 13 |
| American Indian/<br>Alaska Native | 19 (2.8) | 6.8 +/- 1.0 | 14 (74) | 1.0 |
| Asian | 97 (14) | 6.0 +/- 2.0 | 79 (81) | 5.0 |
| Native Hawaiian/Other<br>Pacific Islander | 2 (0.20) | 5.5 +/- 1.0 | 1 (50) | 0.0 |
| White | 487 (73) | 5.9 +/- 2.0 | 329 (68) | 73 |
| Ethnicity |  |  |  |  |
| Hispanic | 68 (10) | 6.2 +/- 1.7 | 46 (68) | 18 |
| Non-Hispanic | 604 (90) | 5.8 +/- 2.0 | 404 (67) | 82 |
| Education |  |  |  |  |
| No High School | 10 (2) | 6.1 +/- 2.5 | 6 (60) | 12 |
| High School | 162 (24) | 5.5 +/- 2.0 | 83 (51) | 27 |
| Some College | 149 (22) | 6.0 +/- 2.0 | 98 (66) | 29 |
| College | 205 (30) | 6.0 +/- 2.0 | 147 (72) | 19 |
| Graduate/Professional | 146 (22) | 6.0 +/- 2.0 | 116 (79) | 12 |
| Employment Status |  |  |  |  |
| Employed | 314 (47) | 5.8 +/- 2.0 | 215 (68) | 51 |
| Unemployed | 194 (29) | 5.7 +/- 2.0 | 111 (57) | 15 |
| Retired | 164 (24) | 6.3 +/- 1.8 | 124 (76) | * |

\*Unable to obtain percentage of U.S. adult population that reports being retired

There were notable geographic differences in COVID-19 vaccine acceptance with DHHS Region 8-Denver having an acceptance rate of over 75% (total sample in region, N = 20; number of participants within sample that would accept COVID-19 vaccine, n = 16) while Region 2-New York (N = 51; n = 22), and Region 5-Chicago (N = 23; n = 9) had an acceptance rate of less than 50% (figure 2a). DHHS Region 8-Denver also had a higher influenza vaccine coverage with over 75% (N = 20; n = 15) of the participants having received the vaccine in the last 8 months, while HHS Regions 2-New York (N = 51; n = 23), 3-Philadelphia (N = 93; n = 44), 6-Dallas (N = 172; n = 81), and 7-Kansas City (N = 18; n = 8) had less than 50% coverage (figure 2b). Figure 3 shows the percent COVID-19 vaccine acceptance against the percent influenza vaccine coverage by state. There was no statistical association between the two variables (coefficient: 0·19; 95% CI: −0·19 – 0·57).

**Figure 2.**
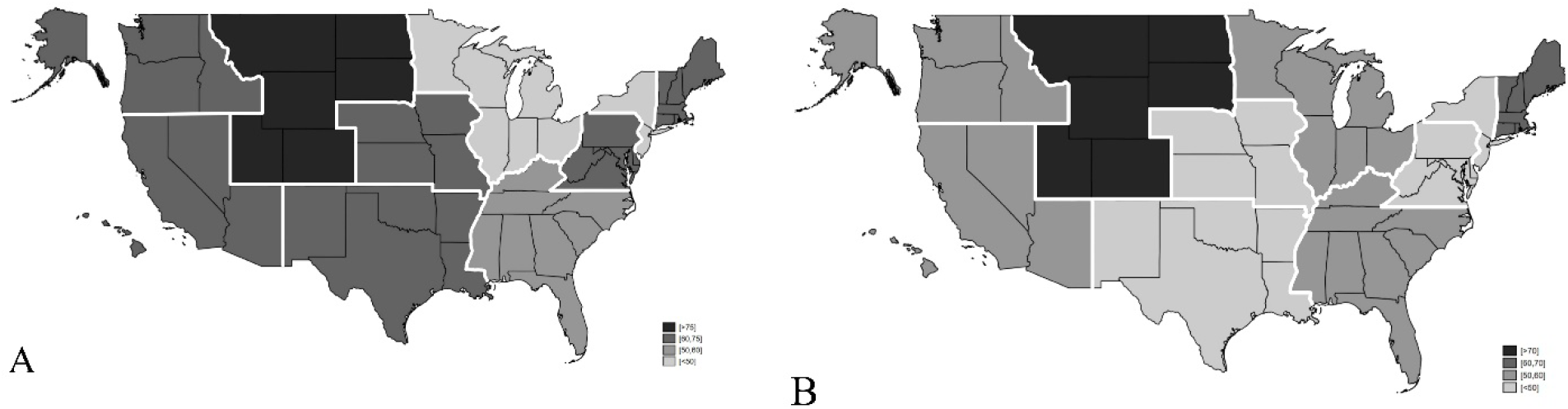
Comparison of COVID-19 vaccine acceptance (A) to reported influenza vaccine uptake (B) in the U.S. by Department of Health and Human Services region.

**Figure 3.**
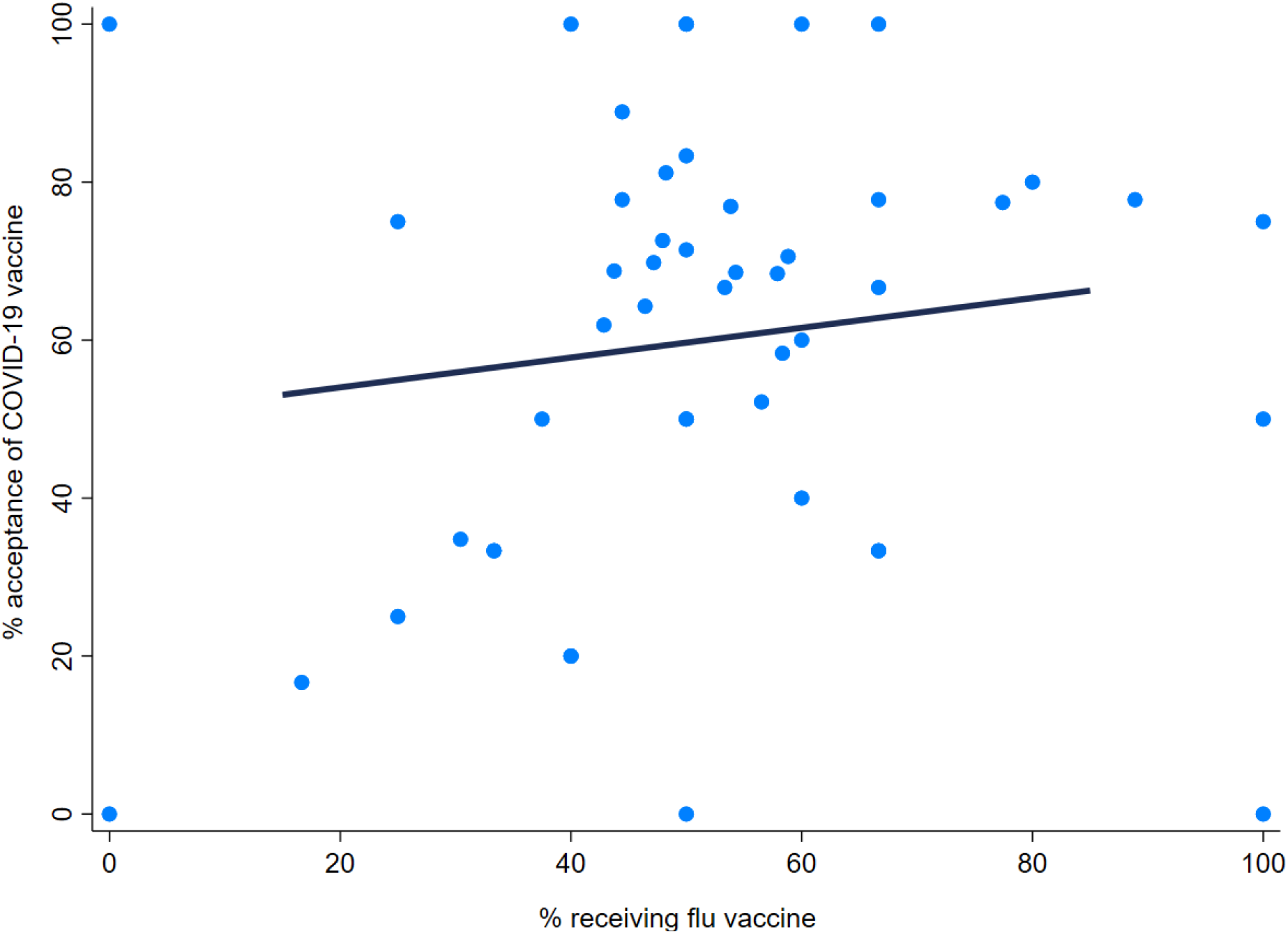
Graph showing the % acceptance of COVID-19 vaccine plotted against the % coverage for influenza vaccine by state. The solid line is the line of best fit using linear regression (coefficient: 0.19; 95% CI: −0.19 - 0.57)

The best model to predict COVID-19 vaccine acceptance in our survey using demographic information that is readily available had age, gender, race, and education as explanatory variables with an area under the curve (AUC) of 72% (table 2; figure 4). Model optimism was estimated to be 1·7% with optimism corrected AUC being 70%.

**Figure 4.**
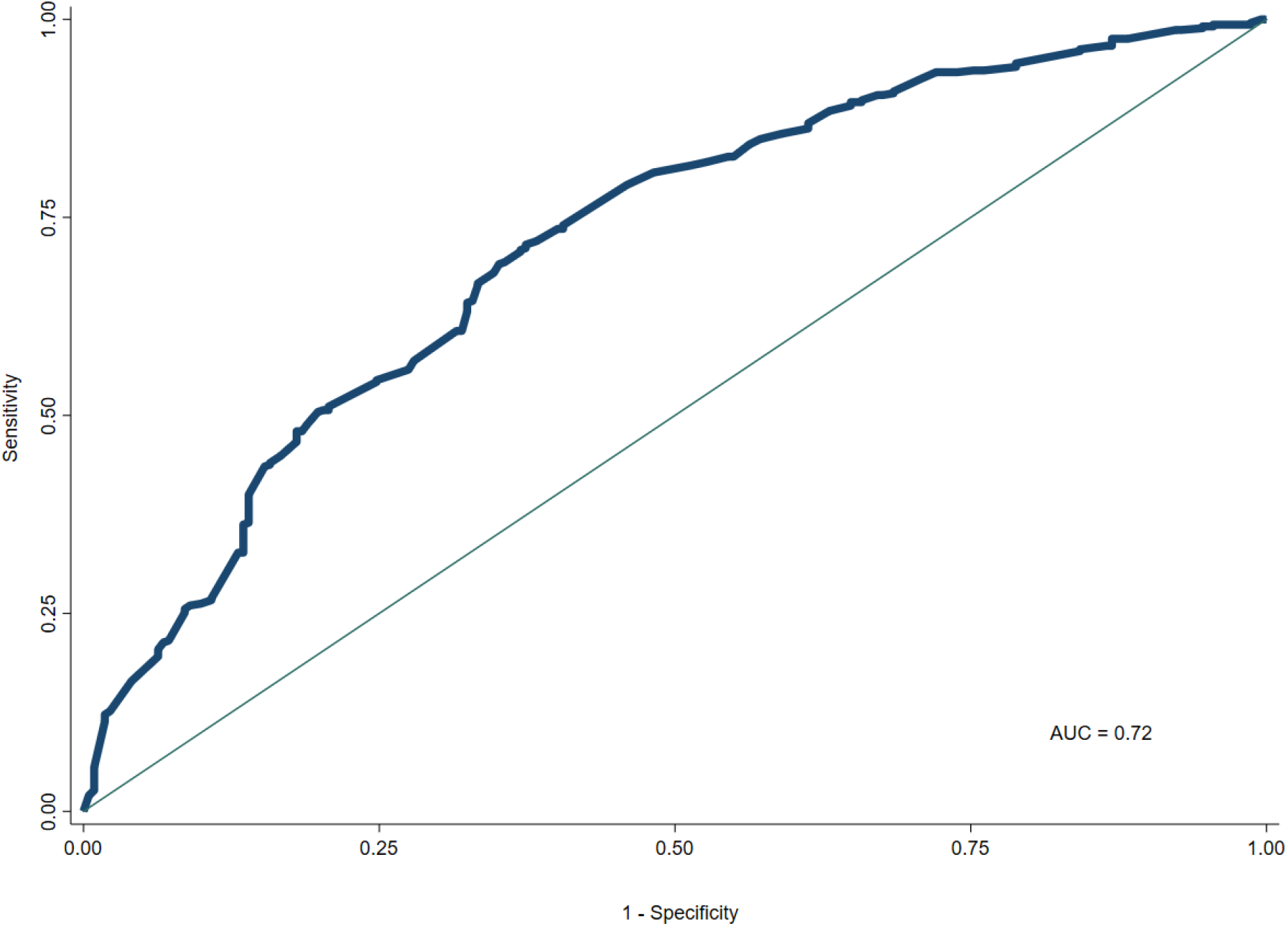
ROC curve for the model logit (COVID-19 vaccine acceptance) = β0 + β1 age (26-35) + β2 age (36-45) + β3 age (46-55) + β4 age (55+) + β5 gender + β6 race (AI) + β7 race (Asian) + β8 race (PI) + β9 race (white) + β10 education (HS) + β11 education (SC) + β12 education (Col) + β13 education (GS)

**Table 2.** Logistic Regression for COVID-19 Vaccine Acceptance by Demographic Characteristics.

|  | OR | SE | CI | *p* |
| --- | --- | --- | --- | --- |
| Gender |  |  |  |  |
| Male | *REF* | *REF* | *REF* |  |
| Female | 0.72 | 0.13 | 0.51 – 1.02 | 0.07 |
| Age (years) |  |  |  |  |
| 18 - 24 | *REF* | *REF* | *REF* |  |
| 25 - 34 | 0.69 | 0.24 | 0.36 – 1.36 | 0.29 |
| 35 - 44 | 0.74 | 0.24 | 0.39 – 1.40 | 0.35 |
| 45 - 54 | 0.55 | 0.19 | 0.28 – 1.08 | 0.08 |
| 55+ | 1.81 | 0.55 | 0.99 – 3.29 | 0.05 |
| Race |  |  |  |  |
| Black/African American | *REF* | *REF* | *REF* |  |
| American Indian/<br>Alaska Native | 4.43 | 2.68 | 1.35 – 14.49 | 0.01 |
| Asian | 6.41 | 2.44 | 3.04 – 13.50 | 0.0001 |
| Native Hawaiian/<br>Pacific Islander | 2.95 | 4.33 | 0.17 – 52.32 | 0.46 |
| White | 3.32 | 0.95 | 1.89 – 5.82 | 0.0001 |
| Education |  |  |  |  |
| No High School | *REF* | *REF* | *REF* |  |
| High School | 0.69 | 0.48 | 0.18 – 2.71 | 0.60 |
| Some College | 1.36 | 0.96 | 0.35 – 5.39 | 0.66 |
| College | 1.78 | 1.25 | 0.45 – 7.03 | 0.41 |
| Graduate/Professional | 2.43 | 1.74 | 0.59 – 9.90 | 0.22 |

The participants reported the highest confidence in healthcare professionals (n = 502; 75%), their own physician (n = 471; 70%), CDC (n = 430; 64%), state health departments (n = 419; 62%), and local health departments (n = 411; 61%). The participants also reported healthcare professionals (n = 503; 75%) and health officials (n = 470, 70%) as the most reliable sources of information on COVID-19. Comparatively, 144 participants (21%) reported social media as a reliable source of COVID-19 information.

## Discussion

While a majority of our respondents from across the US would accept a COVID-19 vaccine, that number may not be sufficient based on some of the estimates COVID-19 herd immunity. With many COVID-19 vaccines under development^11^ and substantial vaccination levels needed to achieve herd immunity, we must clearly understand the hesitancy and acceptance of a COVID-19 vaccine to develop evidenced based interventions. This will allow healthcare professionals and health officials to develop messaging to best address concerns and educate all Americans, especially vulnerable groups.

Our study shows that COVID-19 vaccine acceptance can be predicted with relatively high accuracy by readily available demographic characteristics. Since the beginning of the COVID-19 pandemic in the United States, it has been clear that low-income and communities of color are at higher risk for infection and death from COVID-19.^12^ In fact, when looking at data by zip code from New York City, the dramatic inequality in COVID-19 infections and deaths is evident.^13^ Not only do the most affected zip codes include communities of color, but there are also significant income disparities, demonstrating the intersectionality between race, socio-economic status (SES), and health outcomes.

The disparate health outcomes related to COVID-19 occur not only in New York City, but also across the U.S.^14^ Owen et al. discuss the racial and ethnic differences in COVID-19 infections and deaths in Chicago, Illinois; Charlotte, North Carolina; Milwaukee, Wisconsin and across the states of Michigan and Louisiana.^14^ Historical oppression and current disparities in care are linked to a mistrust of the healthcare system among some Black Americans and may result in these differences in health outcomes.^12^ Supporting this, our study found that Black Americans were less likely to get the influenza vaccine and are less likely to accept a potential COVID-19 vaccine. In addition to racial disparities, COVID-19 vaccine acceptance differs based on education and employment. According to the U.S. Bureau of Labor Statistics, as years of education increases, unemployment rates decrease, and income increases.^15^ Related to this, our study found that as years of education increases, so does reported acceptance of the COVID-19 vaccine. Additionally, unemployed participants reported a lower acceptance rate of a COVID-19 vaccine. These findings demonstrate that low income communities, which are disproportionately impacted by COVID-19,^13^ may be more susceptible to continued outbreaks, even if a vaccine is available.

We need to use caution assuming that reported acceptance or intent translates into actual behavior. This is especially a concern when there is some time between the measurement of intention and the observation of behavior,^16^ which cannot occur until a COVID-19 vaccine is available publicly. Currently, the COVID-19 pandemic is covered on the 24-hour news networks and dominates a great deal of online media. This media coverage may make the COVID-19 pandemic more salient in daily life, especially when compared to influenza. Additionally, during a pandemic and immediately around a new vaccine release, excitement about a vaccine is at its highest.^17^ Another factor that could change salience is if a definitive pharmacological treatment is discovered that reduces duration of illness or deaths. Aside from salience, there may be other unidentified factors that influence COVID-19 vaccine acceptance and eventually vaccine uptake. For example, we found that HHS Region 2-New York, which includes the epicenter for the COVID-19 pandemic in the U.S., had the lowest reported COVID-19 vaccine acceptance. Therefore, COVID-19 vaccine acceptance and eventual uptake may be influenced by more than just media coverage and direct exposure to the economic and health consequences. This means planning for a COVID-19 vaccine should be comprehensive, with a focus on groups that are at high risk.

Building confidence in a COVID-19 vaccine is essential because the herd immunity threshold for SARS-CoV-2, the virus causing COVID-19, is estimated to be between 55% and 82%,^18^ and we found that 67% of our sample would accept the vaccine. Also, the number of Americans who actually receive a COVID-19 vaccine could be lower than those who claim they intend to vaccinate. Thus, health professionals must be careful to encourage trust in vaccination and minimize misinformation. Currently, opposition to vaccination overall may amplify outbreaks^19,20^ like it did during the 2019 measles outbreak. Opposition to vaccines, which occurs actively online,^21^ may influence COVID-19 vaccine acceptance. However, according to Larson, governments that deliberately release reassuring misinformation about COVID-19 may also reduce COVID-19 vaccine acceptance.^20^ In the United States, misinformation released by the government includes the country’s testing capacity,^20^ the efficacy and safety of potential pharmacological interventions,^22^ and the speed at which a vaccine can safely be developed and produced.^23^

To counter this misinformation and improve trust, thoughtful and targeted messaging needs to be developed and tested now to build on the current public interest and continue the momentum past the release of a vaccine. Messaging and education should focus the general American population as well as high risk groups, including low-income individuals and communities of color. This emphasis on high risk communities indicates a need for cultural humility and community engagement. Additionally, how these messages are made available to the public should be considered. We found that our participants had the most trust in COVID-19 information from healthcare professionals and health officials; participants indicated that information from these sources are more reliable than social media. Hence, health officials and healthcare professionals, including nurses and ancillary healthcare staff, should be engaged in community messaging to improve trust in a COVID-19 vaccine and increase uptake.

Our findings may be influenced by possible selection bias because participants needed a CloudResearch account and access to smartphone/computer to participate. Although we had a response rate of 33%, our data are fairly representative of the U.S. population. In addition to the representative sample, strengths of our study include timeliness and ability to stratify on demographic and geographic factors to predict COVID-19 vaccine acceptance. Most importantly, this is one of the first studies that looks at detailed COVID-19 vaccine acceptance.

## Conclusion

Our study found 67% of our sample from across the U.S. would accept a COVID-19 vaccine. However, there were demographic and geographical variations in rates of acceptance that need to be carefully addressed. Policymakers and stakeholders should focus on evidence-based community messaging to improve uptake and break the transmission dynamics.

## Data Availability

Data will be available on request from the corresponding author.

## Author’s contribution

SMM, AAM, and SBO conceptualized the study. SMM, and AAM collected data under supervision from SBO. SMM, AAM, and JE performed and reviewed the analysis, and wrote the initial draft of the manuscript. All authors helped interpret the findings, read and approved the final version of the manuscript.

## Research in context

### Evidence before the study

There are non-peer reviewed surveys suggesting that 75% of the U.S. population would get vaccinated against COVID-19, with 30% getting vaccinated soon after the vaccine is available. Experience from the influenza vaccines and others shows that vaccine uptake is not optimal. Although intent is high, intent does not always translate into behavior.

### Added value of this study

We demonstrate demographic and geographic variations in vaccine intent for COVID-19 with no relationship with the influenza vaccine. We also developed a predictive model to predict COVID-19 vaccine acceptance by readily available demographic information. We found high level of confidence in COVID-19 information received from healthcare professionals and health officials.

### Implications of all the available evidence

Targeted evidenced based community messaging through healthcare professionals and health officials will be required to increase COVID-19 vaccine acceptance.

